# Combined Anticoagulant and antiplatelet therapy is associated with an improved outcome in hospitalized COVID-19 patients: a propensity matched cohort study

**DOI:** 10.1101/2021.07.13.21260467

**Authors:** Kamal Matli, Nibal Chamoun, Aya Fares, Victor Zibara, Soad Al Osta, Rabih Nasrallah, Pascale Salameh, Jacques Mokhbat, Georges Ghanem

## Abstract

**Background:** COVID-19 is a respiratory disease that results in a prothrombotic state manifesting as thrombotic, microthrombotic and thromboembolic events. As a result, several antithrombotic modalities have been implicated in the treatment of this disease. This study aimed to identify if therapeutic anticoagulation or concurrent use of antiplatelet and anticoagulants was associated with an improved outcome in this patient population.

**Methods:** A retrospective observational cohort study of adult patients admitted to a single university hospital for COVID-19 infection was performed. The primary outcome was a composite of in-hospital mortality, ICU admission, or the need for mechanical ventilation. The secondary outcomes were each of the components of the primary outcome, in-hospital mortality, ICU admission, or the need for mechanical ventilation.

**Results:** 242 patients were included in the study and divided into 4 subgroups: therapeutic anticoagulation (TAC), prophylactic anticoagulation + antiplatelet (PACAP), therapeutic anticoagulation + antiplatelet (TACAP), and prophylactic anticoagulation (PAC) which was the reference for comparison. Multivariable cox regression analysis and propensity matching were done and showed when compared to PAC, TACAP and TAC were associated with less in-hospital all cause mortality with an adjusted hazard ratio (aHR) of 0.113 (95% confidence interval (CI) 0.028-0.449) and 0.126 (95% CI, 0.028-0.528) respectively. The number needed to treat (NNT) in both subgroups was 11. Furthermore, PACAP was associated with a reduced risk of invasive mechanical ventilation with an aHR of 0.07 (95% CI, 0.014-0.351). However, the was no statistically significant difference in the occurrence of major or minor bleeds, ICU admission, or the composite outcome of in-hospital mortality, ICU admission or the need for mechanical ventilation.

**Conclusion:** The use of combined anticoagulant and antiplatelet agents or therapeutic anticoagulation alone in hospitalized COVID-19 patients was associated with a better outcome in comparison to prophylactic anticoagulation alone without an increase in the risk of major and minor bleeds. Sufficiently powered randomized controlled trials are needed to further evaluate the safety and efficacy of combining antiplatelet and anticoagulants agents or using therapeutic anticoagulation in the management of patients with COVID-19 infection.

**What is already known about this subject ?:** Covid-19 infection is associated with several complex coagulation disorders resulting in thrombotic, microthrombotic and thromboembolic events. Currently, prophylactic dose anticoagulation is considered the standard of care antithrombotic regimen in hospitalized COVID-19 patients. However, high-quality data about the subject is unavailable.

**What does this study add?:** This is the first adequately sized study in the literature to dwell on the antithrombotic strategy consisting of combination anticoagulant and antiplatelet therapy in the treatment of COVID-19 induced hypercoagulable state. Furthermore, it also challenges the currently recommended prophylactic dosing of anticoagulation used in the treatment of those patients.

**How might this impact on clinical practice?:** Our data suggests for the first time that concurrent use of anticoagulant and antiplatelet therapy is associated with a superior clinical outcome as compared to prophylactic anticoagulation used alone. Furthermore, it solidifies the emerging evidence that therapeutic anticoagulation is linked to better clinical results than prophylactic anticoagulation.

## Introduction

Severe acute respiratory syndrome coronavirus-2 (SARS-CoV-2) has infected over 184 million people and caused over 3.9 million deaths worldwide according to the latest report on the 5^th^ of July by the World Health Organization (WHO).^1^ Although the respiratory symptoms are the primary clinical manifestations of the disease, patients may experience thrombotic complications associated with increased mortality. ^2-4^ COVID-19 also increases cardiovascular disease (CVD), such as myocardial injury, acute coronary syndrome, in addition to venous and arterial thromboembolic events, such as pulmonary embolism, deep venous thrombosis, arterial thrombosis, catheter thrombosis, and disseminated intravascular coagulopathy. ^5–10^

Evidence of hypercoagulability has been observed in markers of coagulation found in COVID-19 patients such as elevated D-dimer and fibrinogen concentration. ^11^ Coagulation in the human body is a complex cascade that involves the interaction between endothelial cells, platelets, and coagulation factors. ^12^ Under normal conditions, platelets circulate in the bloodstream without adhering to the intact and inactive endothelium and most of the clotting factors circulate in an inactive form. ^13^ However, COVID-19 infection was shown to be highly associated with endothelial dysfunction favoring a pro-inflammatory and procoagulant state. ^14^ Infection with this virus leads to subsequent endothelial activation and dysfunction due to disruption of the vascular integrity, leading to endothelial cell apoptosis. This exposes the thrombogenic basement membrane into the circulation and activates the clotting cascade by displaying Von Willebrand Factor (vWF), P-selectin, and fibrinogen, onto which activated platelets bind and play their primary role in thrombosis. ^15^

In addition to the platelet clotting activation, the coagulation cascade is also activated in COVID-19 infected patients. ^16^ This can occur via 2 mechanisms. ^15^ The first mechanism is through the activated platelets that produce Vascular endothelial growth factor (VEGF), which induces endothelial cells to express tissue factor (TF), the main activator of the coagulation cascade. The 2^nd^ mechanism of activation occurs as a direct result of virus induced vessel injury. This is translated clinically into heightened coagulopathy that manifests as microvascular, venous and arterial thrombosis. ^17-19^

Different treatment modalities have been implicated in the treatment of COVID-19 hypercoagulable state with the best agent still undefined. Current guidelines recommend the use of prophylactic dose anticoagulation in all patients hospitalized with COVID-19 infection. ^20,21^ However these recommendations are based on low certainty evidence.

Ongoing clinical trials aim to evaluate the effect of prophylactic and therapeutic anticoagulation therapy on survival and adverse events.^22^ Preliminary data on anticoagulant therapy shows that it appears to be associated with better outcomes and reduced mortality ^23,24^ Furthermore, the role of aspirin was also investigated in COVID-19 patients in a retrospective observational cohort study of adult patients. It was found to be associated with decreased risk of mechanical ventilation, intensive care unit (ICU) admissions, and in hospital mortality after adjusting for confounders. ^25,26^

In this study we evaluated whether the combination of anticoagulation and antiplatelet therapy or therapeutic anticoagulation in hospitalized COVID-19 patients is associated with an improved clinical outcome compared to the standard prophylactic anticoagulation therapy alone as currently recommended by the guidelines. ^27^

## Methods

### Study Settings and Population

Patients with the diagnosis of SARS-CoV-2 infection, admitted to the Lebanese American University-Rizk Hospital between April 2020 and January 31, 2021, and hospitalized in medical wards or ICU, were included in the study. Patients were included if they were aged 18 years and older with confirmed laboratory diagnosis for Covid-19. In accordance with WHO criteria, confirmed cases of SARS-CoV-2 infections were determined by positive results from real-time reverse transcriptase-polymerase chain reaction (RT-PCR) that amplifies DNA sequences specific to the virus from either combination of nasal and pharyngeal swabs or lower respiratory tract aspirates.^1^ Patients on dual antiplatelet therapy, with an acute venous thromboembolism (VTE) defined as deep venous thrombosis (DVT) or pulmonary embolism (PE), acute cardiovascular event, acute stroke (ischemic or hemorrhagic) all within the prior 3 months, or with an active major bleeding, severe thrombocytopenia (< 25,000/mm3), were excluded from this study.

### Institutional Review Board Approval

This study was reviewed and approved by the Lebanese American University Institutional Review Board (IRB) (LAUMCRH.GG4.29/Jan/2021). The study was conducted in accordance with the ethical principles stated in the 1964 Declaration of Helsinki and its amendments.

### Data Collection

All data were collected and screened by local investigators with access to electronic medical records. The patients’ baseline information included demographic characteristics, comorbidities, any known allergies, and chronic medications. Anticoagulation and antiplatelet therapy on admission was collected, as well as its respective indication. Laboratory results and vital signs were recorded on admission. Clinical parameters, including type of oxygen therapy were recorded from admission and up until discharge. Data on complications during the hospital stay, COVID-19 specific pharmacological therapies, and clinical outcome were collected during hospitalization. The data collection sheet was designed in accordance with the toolkit for the collection of thrombosis-related data elements in COVID-19 clinical studies. ^28^ Major bleeding and clinically relevant non-major bleeding were defined as per the International Society of Thrombosis and Haemostasis (ISTH) and Scientific and Standardization Committee (SCC). Major bleeding was bleeding that led to a hemoglobin drop of more than 2 units mg/dL, required more than 2 units of packed red blood cell (RBC) transfusion, lead to death, intracranial bleeding, retroperitoneal bleeding, intraocular bleeding, intra-articular bleeding, pericardial bleeding, spinal bleeding, or intramuscular with compartment syndrome. Clinically relevant non major bleeding was defined as any sign or symptom of bleeding that does not fit the criteria for the ISTH definition of major bleeding but does meet at least one of the following criteria 1) requiring medical intervention by a healthcare professional 2) leading to hospitalization or increased level of care 3) prompting a face to face evaluation. ^29,30^ Minor bleeding was defined as any bleeding Sign or symptom of hemorrhage that doesn’t fit the definitions of major bleed and clinically relevant non-major bleed requiring just telephone or electronic communication without needing to see the physician. Criteria for ICU admission was defined as per the the FDA as having respiratory failure necessitating invasive or non-invasive mechanical ventilation, shock or multiorgan dysfunction/failure in critically ill covid-19. ^31^ Other recorded outcomes related to thrombosis were DVT, PE, stroke, myocardial infarction, and peripheral arterial and other arterial thromboses.

### Antithrombotic Use Definition

At our center, the use of antithrombotic therapy in COVID-19 patients was based on international and institution specific guidelines. ^27^ Prophylactic anticoagulation was the standard of care. Agents used were low molecular weight heparin, unfractionated heparin, fondaparinux or direct oral anticoagulants. Patient with on oral vitamin K antagonists were switched to one of the aforementioned medications to decrease drug-drug interaction risk. Therapeutic dose anticoagulation was chosen when there was an alternative indication for therapeutic anticoagulation or when the patient was considered at high risk/suspicion of thrombosis without documented evidence due to the limited testing as a result of the COVID-19 situation. D-dimer more than 3 times the upper limit of normal, worsening oxygen requirements and sudden deterioration were also used as surrogate markers for the use of therapeutic anticoagulation. Antiplatelet therapy, which consisted of either aspirin or clopidogrel, was added to anticoagulation when the patient was considered at higher risk for CAD based on past medical and social history, age and clinical presentation or was already taking it at home for an alternative indication.

As such, antithrombotic therapy was divided into 4 subgroups: therapeutic anticoagulation (TAC), prophylactic anticoagulation + antiplatelet (PACAP), therapeutic anticoagulation + antiplatelet (TACAP), and prophylactic anticoagulation (PAC) which was used as the reference for comparison with the other subgroups.

### Outcomes

The time from diagnosis to in-hospital mortality, ICU admission, or the need for invasive mechanical ventilation (invasive mechanical ventilation with an endotracheal or tracheostomy tube) were used as the composite primary outcome. The secondary outcomes consisted of the individual events: in-hospital mortality, the need for ICU admissions, and the need for invasive mechanical Ventilation. The outcomes were collected from the patient’s medical records.

### Minimal sample size calculation

Given an alpha of 5% and 80% power with 9 independent variables in the Cox-proportional hazards model, a minimum of 226 patients would be necessary to fit a parsimonious model adjusting for confounding variables; this minimal sample size was calculated using the G*Power software.

### Statistical Analysis

Data analysis was performed using SPSS. Bivariate analysis statistics were conducted among treatment subgroups, using Student test (or Mann-Whitney in case of non-normality or non-homogeneous variances) to compare continuous variables between two subgroups, or Kruskal Wallis test between three subgroups or more, while the Chi-square test (or Fischer’s exact if expected count was lower than 5) was used for categorical variables. In all cases, a p-value lower than 0.05 was considered significant. Imputation of missing variables for CRP, D-dimers and interleukin 6 was used based on the mean. Furthermore, stratification by critical status was not possible, given the small sample size of subgroups.

### Survival analysis

Survival analysis was performed to compare the incidence of the composite outcome and individual outcomes between different treatment exposure subgroups, using the Cox regression modeling. For the purpose of survival analysis, the start of the study was taken as admission to the hospital. The endpoint was defined as the composite outcome, the death of the patient during the hospitalization, being placed on invasive mechanical ventilation, being admitted to the ICU, or being discharged alive from the hospital. Patients still hospitalized as of December 2020 were not included for data collection; only patients who were discharged from the hospital or who died during the study period were considered until the end of the study. Survival time in days was calculated as the difference between the date of hospital admission and the date of event occurrence (ICU admission, in-hospital death, mechanical ventilation, composite outcome, or discharge from the hospital). Log-log plots and Schoenfeld residuals tested the proportional hazards assumption; there were no violations of the proportionality of hazards assumption.

For the composite and each individual outcome, Cox regressions using a backward method. The model variables were selected on the basis of biological plausibility, previously established in the literature and variables with a p-value of 0.2 or less in the bivariate analysis. It included the following variables within the analysis, varying according to the model: anticoagulation and antiplatelet regimen, age, smoker, weight, gender, hypertension, dyslipidemia, diabetes, coronary artery disease, anticoagulation on admission, immunosuppressant, antiplatelet use on admission, inpatient prescription of any of the following medication, azithromycin, fondaparinux, remdesivir, tocilizumab, steroids, tofacitinib oxygen saturation on admission, D-dimers, CRP and interleukin 6, in addition to propensity scores (described below). The Enter method was used to force both propensity scores within the models, in case they were removed by the backward analysis. The regression was then used to estimate adjusted hazard ratios (aHR and their 95% confidence intervals (CIs) for associations between treatment exposure status and the composite outcome and each individual outcome in separate models.

### Propensity scales calculation

To consider factors driving physicians to prescribe anticoagulants and antiplatelet agents in an observational study setting, propensity scales to predict the prescription patterns for both drugs were calculated. The following variables were selected based on known risk factors affecting patient outcomes affecting antithrombotic therapy in accordance with input from the physicians prescribing the treatment strategies: age, gender, smoking status, body weight, admission heart rate, admission temperature, oxygen saturation on admission, hypertension, dyslipidemia, congestive heart failure, diabetes, history of bleeding, liver disease, kidney disease, COPD, coronary artery disease, previous VTE, platelet count, D-dimer, CRP, troponin, fibrinogen, interleukin 6 and chronic anticoagulation prior to admission were included in a backward stepwise logistic regression. The probability of prescription, automatically generated by the software, was used as the propensity score. **(Supplementary material)**

## Results

### Patients Characteristics and Outcomes

Out of 363 COVID-19 patients admitted since April 2020, and after applying eligibility criteria and excluding participants with missing information, a total of 242 patients were included for analysis. Demographic and clinical characteristics by treatment exposure status are outlined in **Table 1**. All patients received anticoagulation therapy, with more than half being on prophylactic anticoagulation therapy versus almost 40% on therapeutic anticoagulation. Antiplatelet therapy was administered to 66.1% of patients, of those, 59.4 % were on PACAP and 40.6% were on TACAP. Upon presentation, 43.4% of patients had dyspnea as a chief complaint. A significant difference between treatment exposure subgroups was detected by oxygen level saturation (p < 0.001) with the lowest oxygen saturation being among patients in the TACAP subgroup. Weight was significantly different among the treatment exposure subgroups, with the average weight being highest in the TACAP subgroup (p=0.039). Moreover, laboratory tests encompassing D-dimer, CRP, and interleukin-6 levels differed between treatment exposure subgroups (p< 0.001, p< 0.001, p= 0.033, respectively), with D-dimer and CRP levels being highest in the TAC subgroup, while interleukin-6 levels being highest in the TACAP subgroup. During their hospital stay, 92.1% of the patients were placed on steroids, 97.5% on vitamin C, 64.9% on vitamin D, and 95.9% received Zinc. Days of oxygen requirement were significantly (P<0.001) different among treatment exposure subgroups with a median of 2,9,10, and 14 days for the PAC, TAC, PACAP respectively.

**Table 1:** Baseline Characteristics and Outcomes by Antithrombotic Treatment Exposure Status.

| Variables | Prophylactic Anticoagulation (PAC)<br>n=51 | Therapeutic Anticoagulation (TAC)<br>n=31 | Prophylactic Anticoagulation+ Antiplatelet Therapy (PACAP)<br>n=95 | Therapeutic Anticoagulation + Antiplatelet Therapy (TACAP)<br>n= 65 | p-Value |
| --- | --- | --- | --- | --- | --- |
| Age, years, mean (SD) | 59.69 (17.04) | 62.55 (15.80) | 66.22 (13.83) | 62.66 (14.73) | 0.09 |
| **Gender** |  |  |  |  |  |
| Males, n (%) | 30 (58.8) | 21 (67.7) | 64 (67.4) | 52 (80.0) | 0.10 |
| Females, n (%) | 21 (41.2) | 10 (32.3) | 31 (32.6) | 13 (20.0) |  |
| Weight (kg), mean (SD) | 78.58 (18.66) | 84.26 (23.91) | 82.02 (17.01) | 88.11 (16.03) | 0.04 |
| Smokers, n (%) | 14 (27.5) | 9 (29.0) | 9 (9.5) | 21 (32.3) | 0.002 |
| Hypertension, n (%) | 20 (39.2) | 14 (45.2) | 55 (57.9) | 33 (50.8) | 0.17 |
| Dyslipidemia, n (%) | 12 (23.5) | 8 (25.8) | 44 (46.3) | 23 (35.4) | 0.03 |
| Congestive Heart Failure, n (%) | 2 (3.9) | 1 (3.2) | 3 (3.2) | 6 (9.2) | 0.32 |
| Cancer, n (%) | 7 (13.7) | 5 (16.1) | 8 (8.4) | 3 (4.6) | 0.21 |
| Diabetes Mellitus, n (%) | 10 (19.6) | 8 (25.8) | 32 (33.7) | 16 (24.6) | 0.29 |
| Bleeding Disorder, n (%) | 1 (2.0) | 0 (0) | 0 (0) | 1 (1.5) | 0.52 |
| Liver Disease, n (%) | 0 (0) | 1 (3.2) | 1 (1.1) | 1 (1.5) | 0.63 |
| Kidney Disease, n (%) | 3 (5.9) | 1 (3.2) | 2 (2.1) | 5 (7.7) | 0.37 |
| COPD, n (%) | 0 (0) | 1 (3.2) | 4 (4.2) | 2 (3.1) | 0.55 |
| CAD, n (%) | 1 (2.0) | 5 (16.1) | 10 (10.5) | 10 (15.4) | 0.09 |
| Others, n (%) | 18 (35.3) | 15 (48.4) | 34 (35.8) | 21 (32.3) | 0.49 |
| Home Anticoagulation, n (%) | 3 (5.9) | 9 (29.0) | 3 (3.2) | 5 (7.7) | <0.001 |
| **Indication for Anticoagulant use** |  |  |  |  |  |
| Atrial Fibrillation, n (%) | 1 (2.0) | 6 (19.4) | 1 (1.1) | 3 (4.6) | 0.004 |
| Venous Thromboembolism, n (%) | 1 (2.0) | 1 (3.2) | 1 (1.1) | 1 (1.5) |  |
| Home Antiplatelet, n (%) | 2 (3.9) | 1 (3.2) | 25 (26.3) | 16 (24.6) | 0.001 |
| **Indication for Antiplatelet use** |  |  |  |  |  |
| Acute Coronary Syndrome, n (%) | 1 (2.0) | 1 (3.2) | 2 (3.2) | 4 (6.2) | 0.06 |
| Post coronary artery bypass grafting, n (%) | 0 (0) | 0 (0) | 5 (5.3) | 2 (3.1) |  |
| Others n (%) | 1 (2.0) | 0 (0) | 15 (15.8) | 7 (10.8) |  |
| ACEi/ARB, n (%) | 8 (15.7) | 9 (29.0) | 29 (30.5) | 20 (30.8) | 0.22 |
| Beta Blocker, n (%) | 13 (25.5) | 8 (25.8) | 23 (24.2) | 19 (29.2) | 0.92 |
| Corticosteroids, n (%) | 3 (5.9) | 1 (3.2) | 1 (1.1) | 0 (0) | 0.13 |
| Antibiotics, n (%) | 1 (2.0) | 0 (0) | 2 (2.1) | 2 (3.1) | 0.81 |
| Immunosuppressant, n (%) | 2 (3.9) | 5 (16.1) | 6 (6.3) | 1 (1.5) | 0.04 |
| **Chief Complaint** |  |  |  |  |  |
| Dyspnea, n (%) | 18 (35.3) | 12 (38.7) | 47 (49.5) | 28 (43.1) | 0.61 |
| Fever, n (%) | 13 (25.5) | 5 (16.1) | 15 (15.8) | 11 (16.9) |  |
| Cough, n (%) | 5 (9.8) | 3 (9.7) | 6 (6.3) | 5 (7.7) |  |
| Desaturation, n (%) | 5 (9.8) | 5 (16.1) | 7 (7.4) | 11 (16.9) |  |
| Vomiting, n (%) | 2 (3.9) | 2 (6.7) | 0 (0) | 2 (3.1) |  |
| Chest Pain, n (%) | 2 (3.9) | 1 (3.2) | 1 (1.1) | 2 (3.1) |  |
| Diarrhea, n (%) | 1 (2.0) | 1 (3.2) | 4 (4.2) | 1 (1.5) |  |
| Fatigue, n (%) | 3 (5.9) | 1 (3.2) | 2 (2.1) | 1 (1.5) |  |
| Chills, n (%) | 0 (0) | 1 (3.2) | 1 (1.1) | 0 (0) |  |
| Myalgia, n (%) | 0 (0) | 0 (0) | 2 (2.1) | 0 (0) |  |
| Others, n (%) | 2 (3.9) | 2 (6.5) | 6 (6.3) | 4 (6.2) |  |
| Days of Symptoms, mean (SD) | 4.92 (4.79) | 4.63 (4.43) | 3.98 (3.82) | 4.19 (3.65) | 0.58 |
| **Oxygen on Presentation** |  |  |  |  |  |
| Nasal Canula, n (%) | 18 (35.3) | 10 (32.3) | 29 (30.5) | 16(24.6) | 0.65 |
| Facemask, n (%) | 2 (3.9) | 4 (12.9) | 2 (2.1) | 6 (9.2) | 0.07 |
| Non-rebreather, n (%) | 2 (3.9) | 3 (9.7) | 4 (4.2) | 13 (20.0) | 0.003 |
| **Vitals on Admission** |  |  |  |  |  |
| Heart Rate, mean (SD) | 90.63 (15.07) | 64.97 (17.28) | 89.56 (13.52) | 92.86 (15.03) | 0.26 |
| Temperature, mean (SD) | 37.70 (1.12) | 38.07 (0.99) | 37.50 (0.90) | 37.74 (1.10) | 0.05 |
| Oxygen Saturation, mean (SD)* | 94.61 (3.20) | 89.74 (10.69) | 93.25 (5.91) | 88.37 (12.73) | 0.001 |
| **Admissions Labs** |  |  |  |  |  |
| Platelet Count, mean (SD) | 265996 (104433) | 262129 (122690) | 239052 (111562) | 227861 (103930) | 0.22 |
| PT, mean (SD) | 17.13 (19.16) | 16.08 (5.13) | 14.06 (2.26) | 14.47 (2.66) | 0.33 |
| PTT, mean (SD) | 31.81 (16.24) | 30.19 (4.46) | 29.03 (5.45) | 32.96 (15.12) | 0.28 |
| D-dimers, mean (SD) | 1.06 (0.81) | 4.60 (7.28) | .90 (0.61) | 1.32 (1.47) | <0.001 |
| **Admissions Labs** |  |  |  |  |  |
| Fibrinogen, mean (SD)* | 484 (141.45) | 524 (214.64) | 511 (131.50) | 532. (171.48) | 0.42 |
| Troponin, mean (SD) | 13.06 (16.34) | 32.69 (49.63) | 25.52 (64.03) | 58.25 (313.92) | 0.59 |
| BNP, mean (SD) | 391.79 (827.93) | 3467.43 (7642.4) | 2078.66 (6510.8) | 1399.73 (3691.12) | 0.48 |
| CRP, mean (SD) | 7.24 (6.01) | 14.92 (9.01) | 6.72 (5.84) | 10.69 (9.56) | <0.001 |
| WBC, mean (SD)* | 11951 (34879) | 8648 (5384) | 6630 (2816) | 7875 (3428) | 0.11 |
| Lymphocyte, mean (SD) | 946 (977.27) | 884 (632.13) | 941 (1186.50) | 662 (380.87) | 0.24 |
| Interleukin-6 Levels, mean (SD) | 85.33 (208.08) | 114.80 (96.301) | 62.07(85.48) | 277.53 (713.05) | 0.03 |
| **VTE Risk (Padua Score)** |  |  |  |  |  |
| Padua Score $\geq 4$, n (%) | 9 (17.6) | 10 (32.3) | 13 (13.7) | 8 (12.3) | 0.07 |
| Padua Score < 4, n (%) | 42 (82.4) | 21 (67.7) | 82 (86.3) | 57 (87.7) |  |
| **Bleeding Risk IMPROVE BLEEDING RISK SCORE** |  |  |  |  |  |
| Score < 7, n (%) | 50 (98.0) | 29 (93.5) | 92 (96.8) | 62 (95.4) | 0.73 |
| Score $\geq 7$, n (%) | 1 (2.0) | 2 (6.5) | 3 (3.2) | 3 (4.6) | |
| **COVID-19 Hospital Administered Medications** |  |  |  |  |  |
| Steroids, n (%) | 42 (82.4) | 31 (100) | 86 (90.5) | 64 (98.5) | 0.004 |
| Tofacitinib, n (%) | 2 (3.9) | 6 (19.4) | 3 (3.2) | 12 (18.5) | 0.001 |
| Remdesivir, n (%) | 12 (23.5) | 12 (38.7) | 19 (20.0) | 32 (49.2) | 0.004 |
| Baricitinib, n (%) | 0 (0) | 1 (3.2) | 3 (3.2) | 1 (1.5) | 0.59 |
| Favipiravir, n (%) | 7 (13.7) | 2 (6.5) | 18 (18.9) | 6 (9.2) | 0.20 |
| Lopinavir-Ritonavir, n (%) | 0 (0) | 1 (3.2) | 1 (0.9) | 0 (0) | 0.08 |
| Tocilizumab, n (%) | 2 (3.9) | 3 (9.7) | 4 (4.2) | 9 (13.8) | 0.09 |
| Ivermectin, n (%) | 1 (2.0) | 2 (6.5) | 1 (1.1) | 0 (0) | 0.13 |
| Vitamin C, n (%) | 49 (96.1) | 29 (93.5) | 94 (98.9) | 64 (98.5) | 0.32 |
| Vitamin D, n (%) | 30 (58.8) | 21 (67.7) | 62 (65.3) | 44 (67.7) | 0.76 |
| Zinc, n (%) | 48 (94.1) | 29 (93.5) | 92 (96.8) | 63 (96.9) | 0.75 |
| Azithromycin, n (%) | 9 (17.6) | 15 (48.4) | 43 (45.3) | 39 (60.0) | <0.001 |
| **Anticoagulant used** |  |  |  |  |  |
| LMWH, n (%) | 47 (92.2) | 26 (83.9) | 77 (81.1) | 55 (84.6) | 0.36 |
| UFH, n (%) | 3 (5.9) | 1 (3.2) | 3 (3.2) | 4 (6.2) | 0.77 |
| Fondaparinux, n (%) | 0 (0) | 2 (6.5) | 12 (12.6) | 7 (10.8) | 0.03 |
| DOACs, n (%) | 0 (0) | 2 (6.5) | 3 (3.2) | 0 (0) | 0.11 |
| **Antiplatelet used** |  |  |  |  |  |
| Aspirin, n (%) | 0 (0) | 0 (0) | 93 (97.9) | 62 (95.4) | < 0.001 |
| Clopidogrel, n (%) | 0 (0) | 0 (0) | 2 (2.1) | 5 (7.7) | 0.05 |
| **Outcomes** |  |  |  |  |  |
| *Composite, n(%) | 8 (15.7) | 9 (29.0) | 12 (12.6) | 34 (52.3) | <0.001 |
| Mortality, n(%) | 5 (9.8) | 7 (22.6) | 6 (6.3) | 16 (24.6) | 0.004 |
| ICU Admission, n(%) | 7 (13.7) | 8 (25.8) | 12 (12.6) | 32 (49.2) | <0.001 |
| Invasive ventilation, n(%) | 4 (ca) | 6 (19.4) | 5 (5.3) | 19 (29.2) | <0.001 |
| **Major Bleeding, n (%)** | 2 (3.9) | 2 (6.5) | 0 (0) | 6 (9.2) | 0.17 |
| **Clinically relevant non-major bleeds, n (%)** | 2 (3.9) | 1 (3.2) | 0 (0) | 6 (9.2) | 0.29 |
| **Minor Bleeds, n (%)** | 1 (2.0) | 0 (0) | 3 (3.2) | 4 (6.2) | 0.39 |
| **Any thromboembolic event, n (%)** | **5 (9.8)** | **9 (38.7)** | **7 (7.4)** | **15 (32.3)** | **0.002** |
| DVT, n (%) | 0 (0) | 0 (0) | 0 (0) | 0 (0) | - |
| PE, n (%) | 0 (0) | 3 (9.7) | 0 (0) | 1 (1.5) | 0.002 |
| MI, n (%) | 0 (0) | 0 (0) | 2 (2.1) | 4 (6.2) | 0.12 |
| Ischemic Stroke, n (%) | 0 (0) | 0 (0) | 0 (0) | 1 (1.5) | 0.43 |
| Unknown Stroke, n (%) | 1 (2.0) | 2 (6.5) | 1 (1.1) | 2 (3.1) | 0.40 |
| Others Arterial Disease, n (%) | 4 (7.8) | 7 (22.6) | 4 (4.2) | 13 (20.0) | 0.003 |

\*Composite comprised of either Mortality, ICU Admissions, or Mechanical Ventilation.
COPD: Chronic Obstructive Pulmonary Disease ;CAD: Coronary Artery Disease ; ACEi /ARB: Angiotensin converting enzyme inhibitor/ Angiotensin Receptor Blocker; BNP: Brain Natriuretic Peptide; CRP: C-Reactive Protein; WBC : White Blood Cells; DVT: Deep Vein Thrombosis; PE: Pulmonary Embolism; VTE: Venous Thromboembolism; PT: Prothrombin Time; PTT: Partial thromboplastin time; MI: Myocardial Infarction; TIA: Transient Ischemic Attack ; ICU: Intensive Care Unit; HFNC: High Flow Nasal Canula; LMWH: Low Molecular Weight Heparin; UFH: Unfractionated Heparin; DOAC: Direct Oral Anticoagulant ; SD: Standard Deviation.

As seen in **Table 1**, composite endpoint that combines mortality, admission to ICU, or need for invasive ventilation was reached in 26% of the sample among which 12.7%, 14.3%, 19.1%, and 53.9% were in the PAC, TAC, PACAP, and TACAP treatment exposure subgroups, respectively. The individual clinical outcomes of mortality, ICU admission, and mechanical ventilation occurred in 14.1%, 24.4%, and 14.1%, respectively of the sample. There was no difference in the rates of bleeding observed in the study. Major bleeding occurred in 10 out 242 patients (4.1%) of the study population with no significant difference across the subgroups (p=0.17). Clinically relevant non-major bleeds occurred in 9 out of 242 patients (3.7%) of the study with no significant difference across the treatment subgroups (p=0.39). Thirty-five patients (14.4%) experienced thrombosis during the study period. There was a significant (p=0.002) difference in the occurrence of thrombosis across the study subgroups, PAC 4 out of51 patients (7.8%), TAC, 9 out of31 patients(29.0%), PACAP 7 out of 95patients (7.4%) and TACAP 15 out of 65patients (23.1%).

### Propensity matched Multivariable Cox regression analysis results

Results of the multivariable Cox regression analysis showed no significant difference amongst the three antithrombotic regimens as compared to prophylactic dose with respect to the composite outcome. Higher oxygen saturation on admission showed to be protective. For every 1 unit increase in oxygen saturation on admission patients had a 4.6% decrease in the risk of the composite. Hypertension or being prescribed tocilizumab were significantly associated with an increased risk of experiencing the composite outcome. Elevated interleukin 6 levels were also associated with a mild but statistically significant increase in the composite (aHR)=1, 95%CI 1.000-1.001, p=0.026). (**Table 2 a**)

**Table 2.**
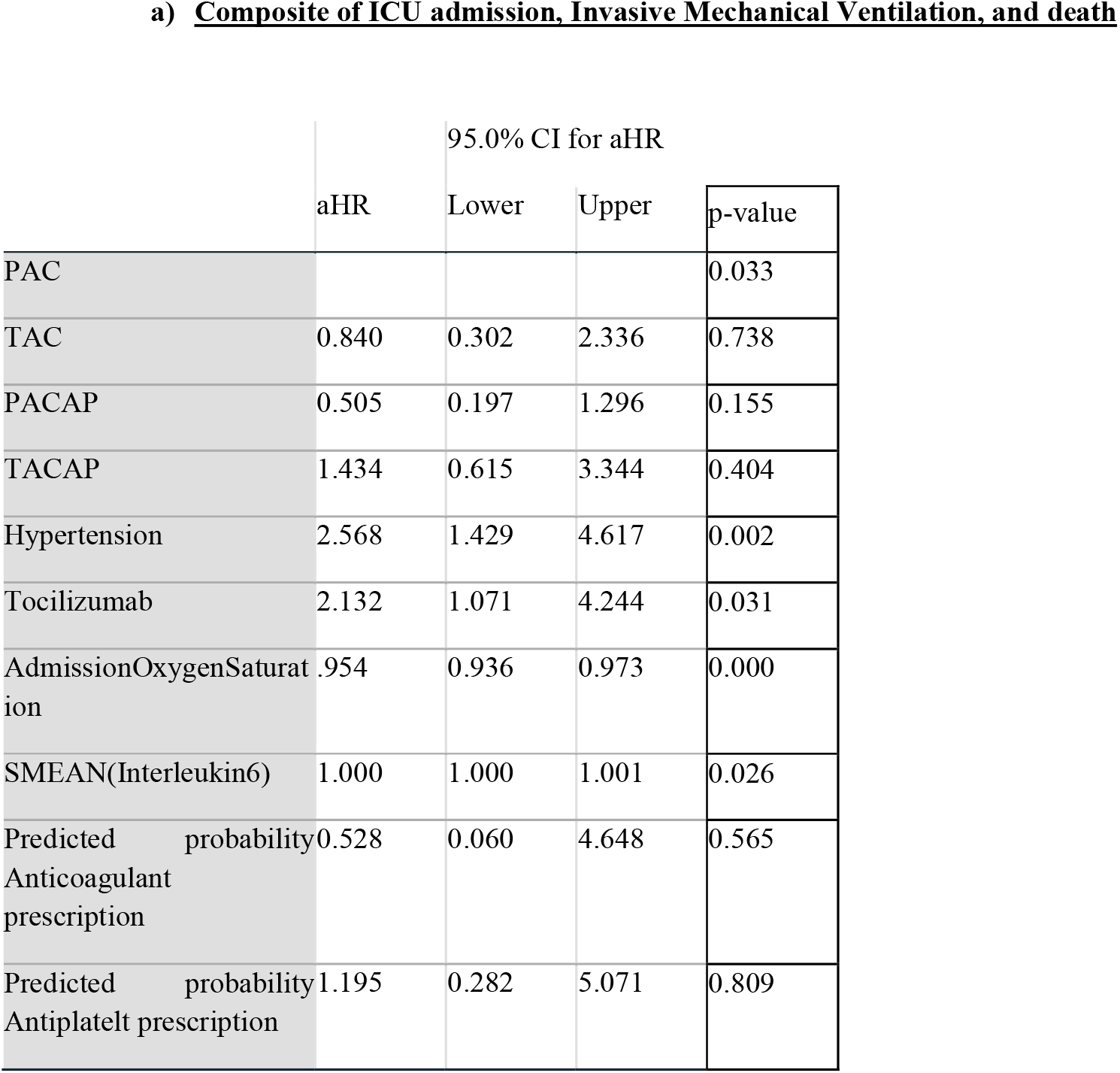
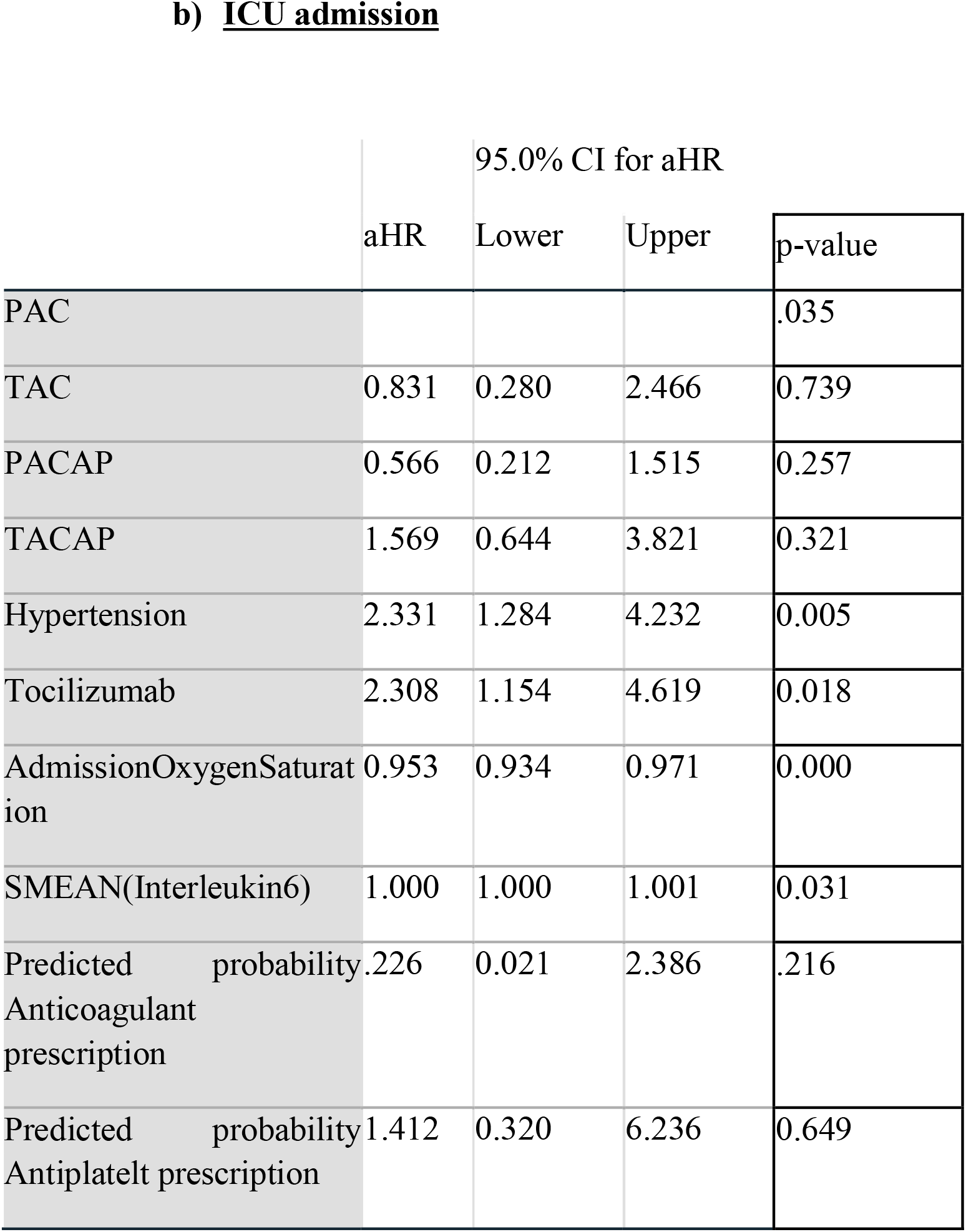
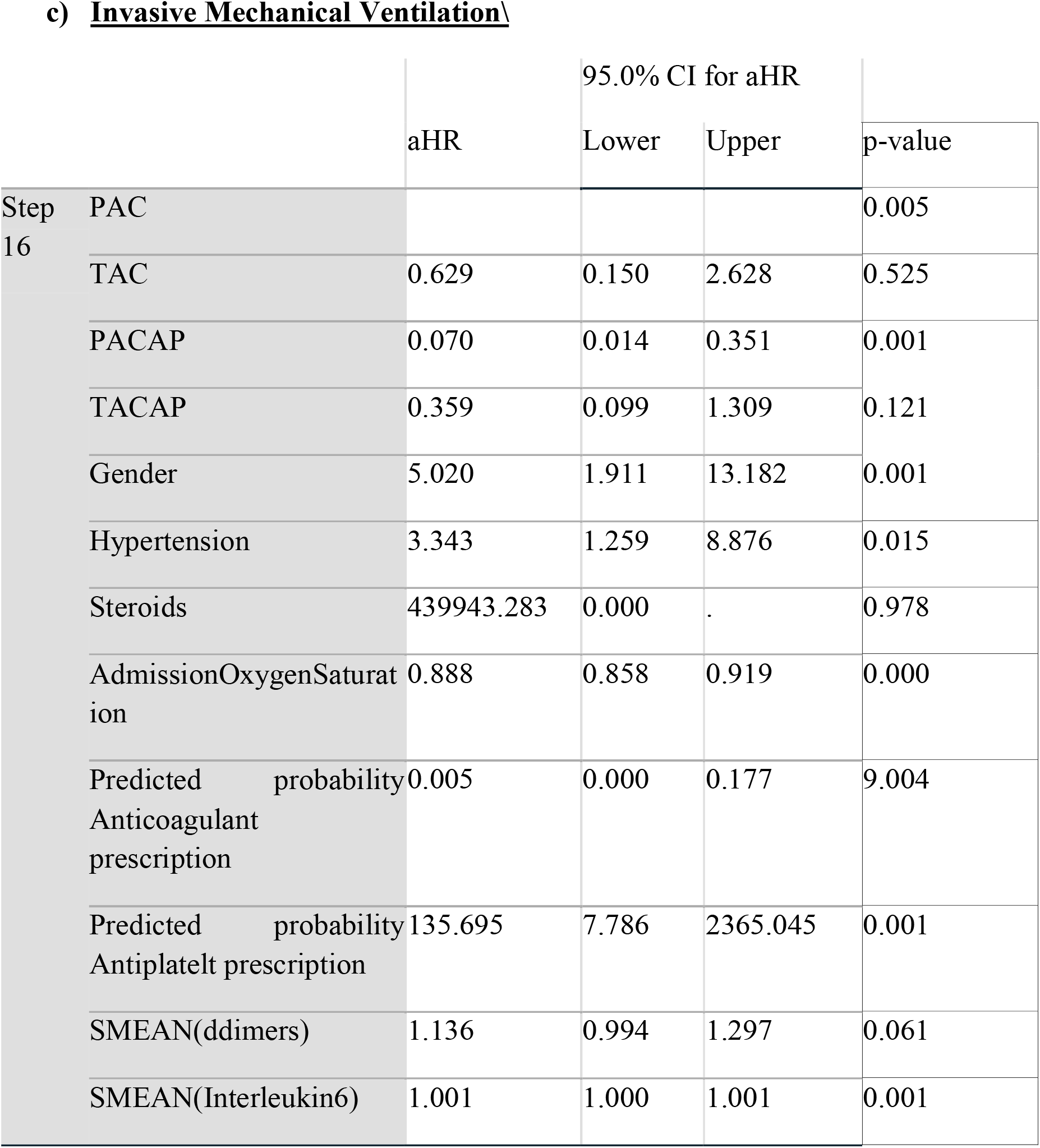
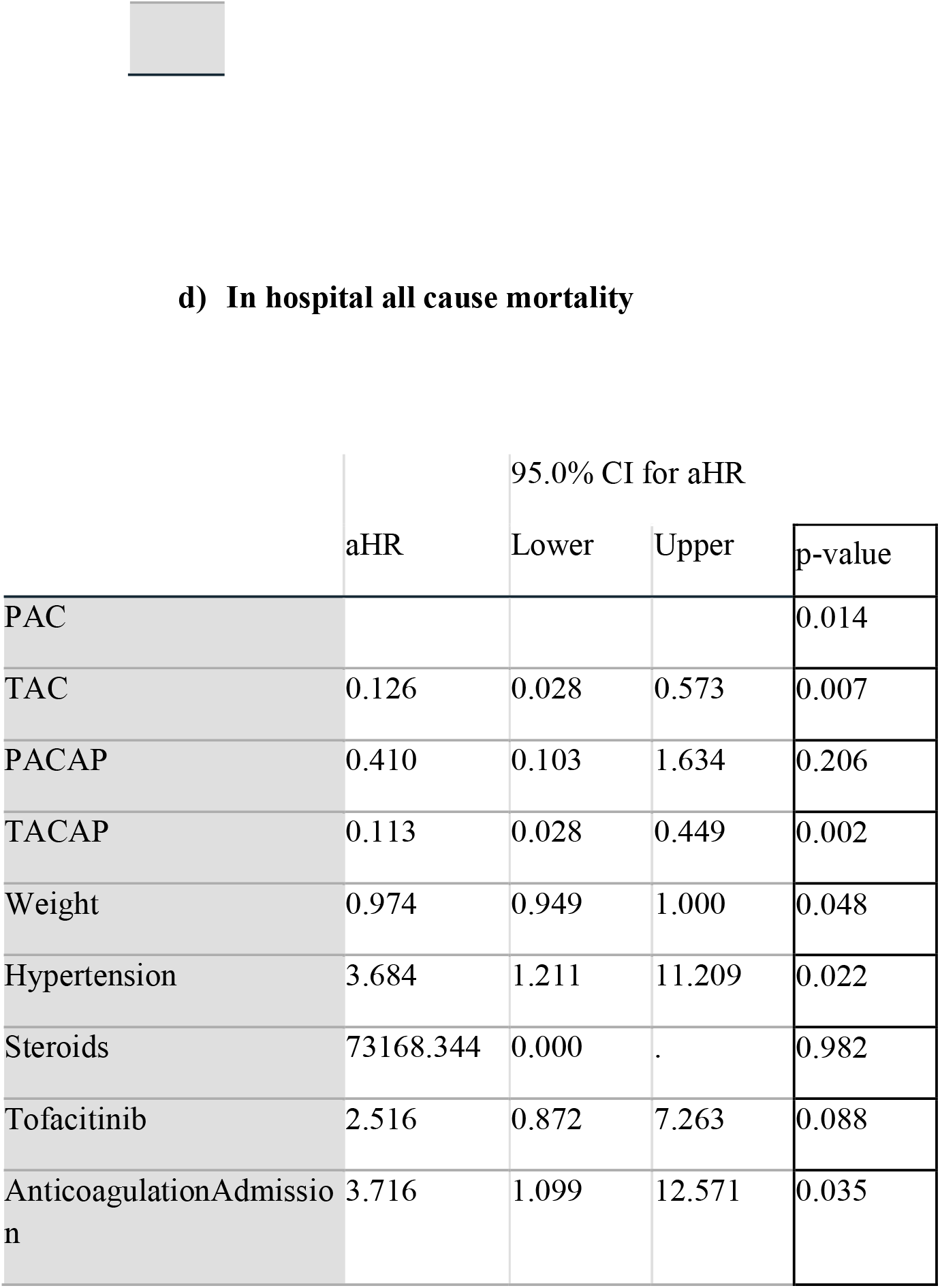
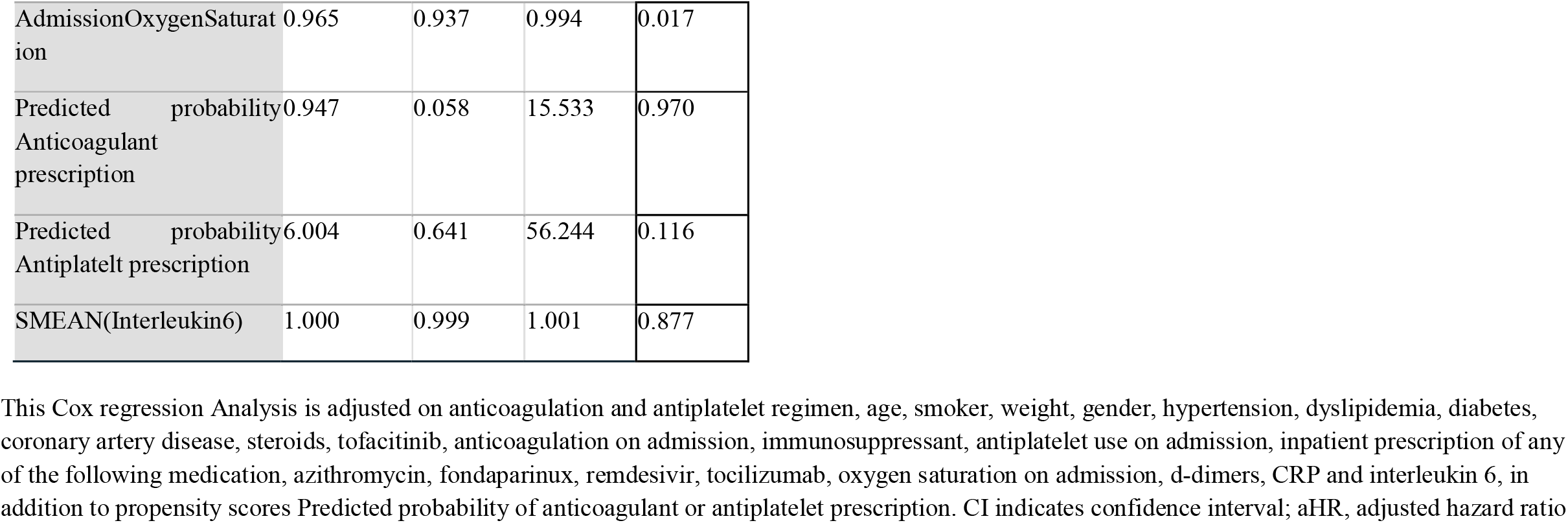
Multivariable Cox regression analysis according to the different antithombotic regimens to define (a) the composite endpoint of ICU admission, Invasice Mechanical Ventilation, and death (b) ICU admission (c) Invasive Mechanical Ventilation (D) In-hospital all cause mortality.

Patients on PACAP had a 93% decreased risk of experiencing invasive mechanical ventilation (aHR=0.070; 95CI=0.014-0.351, P=0.001). For every unit increase in oxygen saturation on admission the risk of invasive mechanical ventilation decreased by 11.2%. **(Table 2 c)** Elevated interleukin 6 levels were also associated with a statistically very mild increase in the composite (aHR=1.001, 95%CI 1.000-1.001, p=0.001). Hypertension and the female sex were associated with 3 to 5 fold increase in the risk of invasive mechanical ventilation respectively **(Table 2 c)**

Therapeutic anticoagulation regimens with or without concurrent antiplatelet use was associated with a 87.4% and 88.7% of decreased risk of mortality respectively. The calculated number needed to treat (NNT) in both subgroups was 11 (**Table 2 d**). However when TACAP was compared to TAC alone, there was no statistically significant difference in the rates of mortality (16 (24.6%) vs 7(22.6%) respectively, p=0.809).**(Supplementary Material)**

Higher bodyweight also showed to be protective. For every 1 kg of bodyweight, patients’ risk of mortality decreased by 2.6%. Every 1 unit increase of oxygen on admission was associated with a 2.6% decrease in mortality. Both hypertension and prior anticoagulation use on presentation were associated with around a 3.7 fold increase in mortality. **(Table 2 d)**

The results showed no significant difference amongst the three antithrombotic regimens as compared to prophylactic dose with respect to the ICU admission rate. Higher oxygen saturation on admission showed to be protective decreasing the risk of ICU admission by 4.7% for every unit of oxygen. Hypertension or being prescribed tocilizumab during hospitalization were significantly associated with around twice the risk of ICU admission. Elevated interleukin 6 levels were also associated with a statistically very mild increase in the composite (aHR=1, 95%CI 1.000-1.001, p=0.031). (**Table 2 b**)

## Discussion

In our retrospective study, a propensity matched multivariable cox regression model demonstrated that the use of concurrent therapeutic anticoagulation and antiplatelet therapy or therapeutic anticoagulation alone was associated with improved outcomes in patients hospitalized for COVID-19 infection when compared to patients receiving the standard of care as per current guidelines which is prophylactic dose anticoagulation. A statistically significant reduction in the rate of all cause in-hospital mortality with no increased rates of major or minor bleeds was found. Furthermore, we demonstrated that prophylactic anticoagulation plus antiplatelet therapy significantly reduced the rates of invasive mechanical ventilation compared to prophylactic anticoagulation alone. Importantly, this is the first adequately sized study that clearly dwells on the use of combined anticoagulation and antiplatelet therapy for the treatment of the COVID-19 induced hypercoagulable state.

Covid-19 has been associated with an increased risk of aterial, venous and microvascular thrombosis secondary to endothelial dysfunction with reported cases of heparin resistance.^15^ As such, it was no surprise that we observed in our study that therapeutic anticoaglution with or without concurrent antiplatelet use was associated with decreased all cause motrality when compared to prophylactic anticoagulation alone. However, after comparing in this study we were unable to discern a difference between therapeutic anticoagulation to therapeutic anticoagulation plus antiplatelet. Our results are in line with existing literature. Roomi et. Al conducted a retrospective cohort consisting of 176 patients admitted to the hospital for COVID-19 infection and reported that therapeutic anticoagulation was associated with a lower rate of in-hospital mortality compared to prophylactic anticoagulation (OR 3.05, 95% CI 1.15–8.10, p = 0.04). ^32^ Pre-print publications by the multiplatform collaborative clinical trial (ACTIV-4a [NCT04505774], REMAP-CAP [NCT02735707], ATTACC [NCT04372589]) that aimed to assess the benefit of full dose anticoagulation to treat non critically-ill or critically ill adults hospitalized for COVID-19, compared to prophylactic dosing showed that therapeutic anticoagulation had a trend toward less mortality in non-critically ill patients. ^33^ Moreover, therapeutic dose anticoagulation has also been shown to reduce endothelial cell lesion (p = 0.02) which could also reduce the thromboembolic risk of COVID-19, suggesting another therapeutic target for anticoagulants. ^34^ Our results are not in line with those of Chocron et.al who reported no association between various doses of anticoagulation during hospitalization for COVID-19 infection with an improved outcome. ^35^ Nevertheless, in the multivariable analysis they performed, they did not account for the use for inpatient treatment agents that have been proven to modify outcomes in COVID-19 patients such as steroids, Remdesivir, and Tocilizumab. ^36-38^ IL-6 levels and D-dimer ^39^, markers predictive of outcomes in COVID-19, were also not accounted for. As such, this increases the risk of a bias in their analysis of association between anticoagulation and clinical outcome.

Prophylactic anticoagulation plus antiplatelet, mainly aspirin, was found to be associated with a statistically significant reduction in the need for mechanical ventilation when compared to prophylactic anticoagulation. This observed benefit is probably due to aspirin’s established role in decreasing inflammation, reducing platelet-neutrophil aggregates in the lungs, and increasing lipoxin formation which restores pulmonary endothelial function. ^40^ Chow et al. demonstrated in a retrospective cohort study that when patients were given aspirin within 24 hours of hospital admission for COVID-19 infection, they had decreased rates of mechanical ventilation (adjusted HR: 0.56; 95% [CI]: 0.37-0.85; p=0.007), ICU admission (adjusted HR: 0.57; 95% [CI]: 0.38-0.85; p=0.005), and in-hospital mortality (adjusted HR 0.53; 95% [CI]: 0.31-0.90; p=0.02) without increase in major bleeding (p=0.69) or overt thrombosis (p=0.82) in comparison to those who did not receive aspirin. ^41^ Furthermore, a meta-analysis by Panka et. al showed that aspirin is effective in preventing and treating ARDS. ^40^ Our results are in line with a small case-control study where five COVID-19 infected patients were placed on fondaparinux and an antiplatelet therapy regimen (aspirin and/or clopidogrel and a continuous infusion of tirofiban), while controls received prophylactic or therapeutic heparin infusion. Treatment in the combination arm hinted towards improved gas exchange, increase in arterial oxygenation and A-a O2 difference in COVID-19 infected patients. ^42^ Yet, therapeutic anticoagulation plus antiplatelet use was not associated with a decrease in mechanical ventilation. This is probably attributed to the fact that the patients with more severe presentations were selected to be given therapeutic anticoagulation which would falsely hide the benefit of aspirin in lieu of their disease severity.. Paranjpe et al. reported an increased rate of mechanical ventilation among COVID-19 patients receiving therapeutic anticoagulation and attributed this finding to the fact that therapeutic anticoagulation is reserved for the sicker patients. To note that after adjusting for mechanical ventilation, they found an improved survival in those patients. ^43^

We hypothesize that the combined effect of antiplatelet and anticoagulant therapy on COVID-19 induced platelet thrombosis and hypercoagulability, respectively, may result in a synergistic and superior outcome than using either agent alone. Especially since the thrombotic manifestations of COVID-19 are heterogenous and arising from different hypercoagulable mechanisms occurring independently or simultaneously.

Several limitations of this study should be mentioned. Given that this is a retrospective cohort study, association and not causality can be reported between the antithrombotic regimens and outcomes. The sample size was relatively small which makes it difficult to completely adjust for confounding and limits generalizability of the results. Despite efforts to control confounders by using different analytical strategies, some potential biases may have been disregarded, leading to a potential residual confounding. Data are based on the experience of a single center in Lebanon which prevents generalizability of our findings based on a potential selection bias. Furthermore, thrombotic events may have been underreported due to the strict isolation measures for covid-19, which may have led treating physicians to underuse imaging for appropriate diagnosis, generating a possible information bias.

Strengths of the study include that it may be one of the first studies to examine the combination of therapies for the management of SARS COV-2 to prevent adverse outcomes. In addition, to adjust the analysis for significant variables a propensity matched multivariable cox regression model was employed. Lastly, hazards models that allow time to event analysis accounting for multiple outcomes were employed.

## Conclusion

In conclusion, this is the first study to demonstrate an improved clinical outcome with the use of combined anticoagulant and antiplatelet therapy in comparison to prophylactic anticoagulation alone in patients hospitalized for covid-19 infection with no subsequent increase in minor or major bleeding risk. Furthermore, also therapeutic anticoagulation was found to be superior prophylactic anticoagulation and associated with a better outcome. Prospective randomized controlled trials are needed for the evaluation of the safety and efficacy of combining antiplatelet and anticoagulants agents in the management of COVID-19 patients and to identify the optimal dosing of anticoagulants.

**Figure 1.**
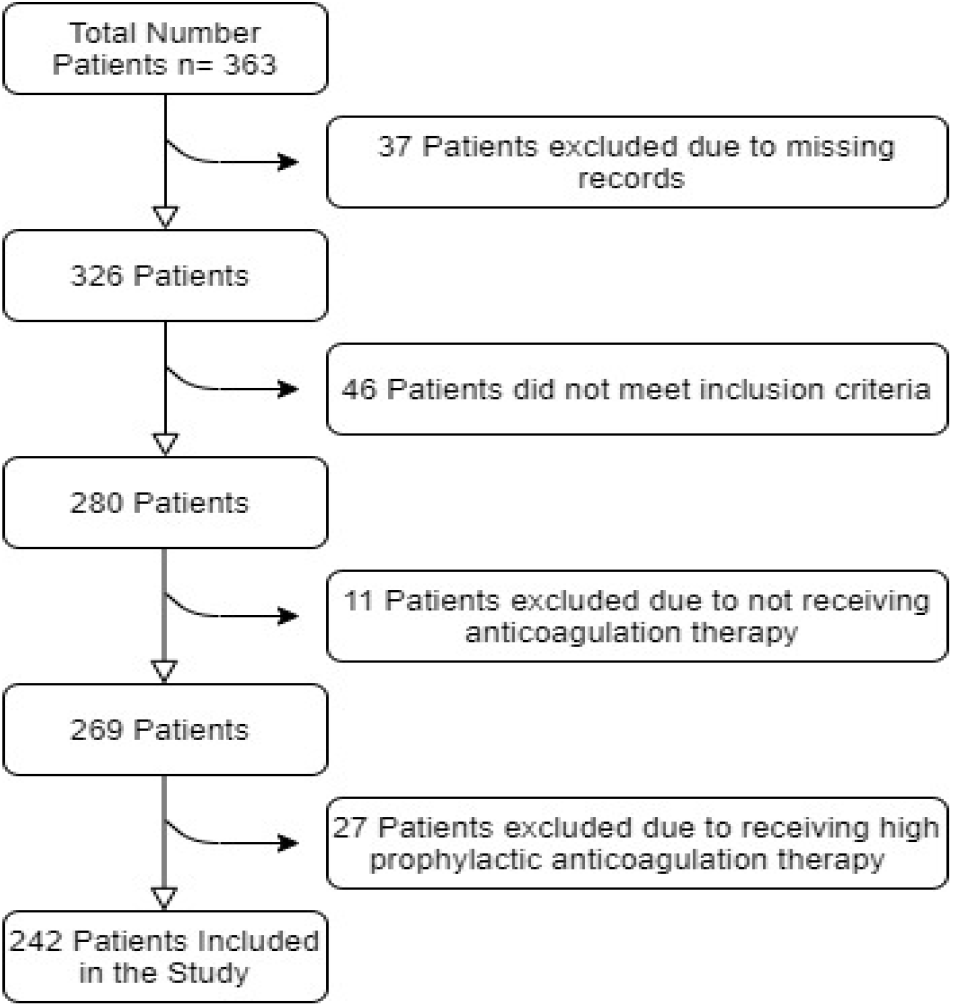
Flowchart of study participants.

**Figure 2.**
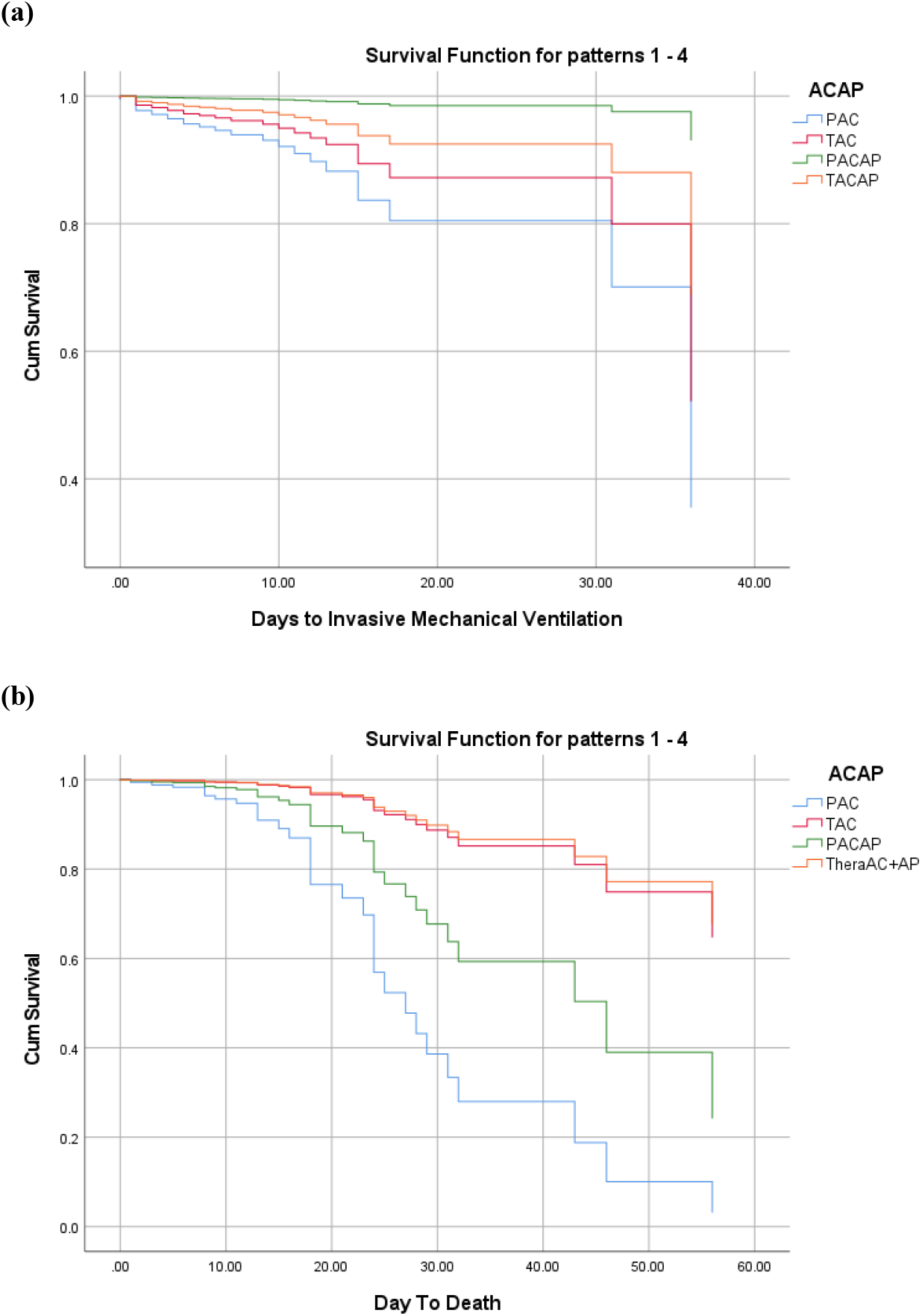
Cox Regression Model for (a) Days to Invasive Mechanical Ventilation (b) Days to Death. Survival function for a) invasive mechanical ventilation b) all cause mortality. Patients are stratified according to antithrombotic regimen used. **a)** PACAP was associated with a decreased hazard for receiving invasive mechanical ventilation (aHR=0.070; 95CI=0.014-0.351). **b)** TACAP and TAC were associated with less in-hospital all cause mortality with an aHR of 0.113 (95% CI 0.028-0.449) and 0.126 (95% CI, 0.028-0.528) respectively. CI indicates confidence interval; aHR, adjusted hazard ratio

## Data Availability

The data is available upon resonable request from the last author

## Acknowledgements

We thank Dr. Christy Costanian for her helpful input concerning statistics and initial study organisation.

## Contributors

All coauthors contributed to this work. KM contributed to hypothesis conception, study design, data analysis and interpretation and wrote the final draft of manuscript. NC contributed to study design, data analysis and interpretation and contributed to writing the manuscript. AF contributed to data extraction and contributed to writing the manuscript. VZ contributed to data entry and analysis and contributed to writing the manuscript. SA and RN contributed to data extraction and managed the references. PS contributed to data analysis and interpretation as well as critical revisions of the manuscript. JM and GG contributed to critical revisions of the manuscript. All authors read and approved the final version of the manuscript before submission for publication.

## Data availability statement

All data relevant to the study are included in the article and are available on reasonable request from the last author GG

## Funding

None

## Competing interests

None

## Patient consent for publication

Not required.

## Notes

### Competing Interest Statement

The authors have declared no competing interest.

### Funding Statement

No funding avaialble

## Referencces

1. World Health Organization. Coronavirus disease (COVID-19) pandemic. 2021 (https://www.who.int/emergencies/diseases/novel-coronavirus-2019.

2. Klok FA, Kruip MJHA, van der Meer NJM, et al. Confirmation of the high cumulative incidence of thrombotic complications in critically ill ICU patients with COVID-19: An updated analysis. Thromb Res. 2020;191:148–150. doi:10.1016/j.thromres.2020.04.041

3. Cui S, Chen S, Li X, Liu S, Wang F. Prevalence of venous thromboembolism in patients with severe novel coronavirus pneumonia. J Thromb Haemost. 2020;18(6):1421–1424. doi:10.1111/jth.14830

4. Middeldorp S, Coppens M, van Haaps TF, et al. Incidence of venous thromboembolism in hospitalized patients with COVID-19. J Thromb Haemost. 2020;18(8):1995–2002. doi:10.1111/jth.14888

5. Barbar S, Noventa F, Rossetto V, et al. A risk assessment model for the identification of hospitalized medical patients at risk for venous thromboembolism: the Padua Prediction Score. J Thromb Haemost. 2010;8(11):2450–2457. doi:10.1111/j.1538-7836.2010.04044.x

6. Huang C, Wang Y, Li X, et al. Clinical features of patients infected with 2019 novel coronavirus in Wuhan, China [published correction appears in Lancet. 2020 Jan 30;:]. Lancet. 2020;395(10223):497–506. doi:10.1016/S0140-6736(20)30183-5

7. Wang D, Hu B, Hu C, et al. Clinical Characteristics of 138 Hospitalized Patients With 2019 Novel Coronavirus-Infected Pneumonia in Wuhan, China [published correction appears in JAMA. 2021 Mar 16;325(11):1113]. JAMA. 2020;323(11):1061–1069. doi:10.1001/jama.2020.1585

8. Zhou F, Yu T, Du R, et al. Clinical course and risk factors for mortality of adult inpatients with COVID-19 in Wuhan, China: a retrospective cohort study [published correction appears in Lancet. 2020 Mar 28;395(10229):1038] [published correction appears in Lancet. 2020 Mar 28;395(10229):1038]. Lancet. 2020;395(10229):1054–1062. doi:10.1016/S0140-6736(20)30566-3

9. Wu Z, McGoogan JM. Characteristics of and Important Lessons From the Coronavirus Disease 2019 (COVID-19) Outbreak in China: Summary of a Report of 72lJ314 Cases From the Chinese Center for Disease Control and Prevention. JAMA. 2020;323(13):1239–1242. doi:10.1001/jama.2020.2648

10. Ruan Q, Yang K, Wang W, Jiang L, Song J. Clinical predictors of mortality due to COVID-19 based on an analysis of data of 150 patients from Wuhan, China [published correction appears in Intensive Care Med. 2020 Apr 6;:]. Intensive Care Med. 2020;46(5):846–848. doi:10.1007/s00134-020-05991-x

11. Spiezia L, Boscolo A, Poletto F, et al. COVID-19-Related Severe Hypercoagulability in Patients Admitted to Intensive Care Unit for Acute Respiratory Failure. Thromb Haemost. 2020;120(6):998–1000. doi:10.1055/s-0040-1710018

12. Transfusion Medicine and Hemostasis - 3rd Edition. https://www.elsevier.com/books/transfusion-medicine-and-hemostasis/shaz/978-0-12-813726-0 (accessed 20 Jan 2021)

13. Palta S, Saroa R, Palta A. Overview of the coagulation system. Indian J Anaesth. 2014;58(5):515–523. doi:10.4103/0019-5049.144643

14. Biswas I, Khan GA. Endothelial dysfunction in cardiovascular diseases. Basic Clin Underst Microcirc. 2020.doi:10.5772/intechopen.89365

15. Matli K, Farah R, Maalouf M, et al. Role of combining anticoagulant and antiplatelet agents in COVID-19 treatment: a rapid review. Open Heart 2021;8:e001628. doi: 10.1136/openhrt-2021-001628

16. Jin Y, Ji W, Yang H, Chen S, Zhang W, Duan G. Endothelial activation and dysfunction in COVID-19: from basic mechanisms to potential therapeutic approaches. Signal Transduct Target Ther. 2020;5(1):293. doi:10.1038/s41392-020-00454-7

17. Magro C, Mulvey JJ, Berlin D, et al. Complement associated microvascular injury and thrombosis in the pathogenesis of severe COVID-19 infection: A report of five cases. Translational Research. 2020;220:1–13. doi: 10.1016/j.trsl.2020.04.007;

18. De Roquetaillade C, Chousterman BG, Tomasoni D, et al. Unusual arterial thrombotic events in Covid-19 patients. Int J Cardiol. 2021;323:281–284. doi:10.1016/j.ijcard.2020.08.103

19. Li W, Xu Z, Xiang H, Zhang C, Guo Y, Xiong J. Risk factors for systemic and venous thromboembolism, mortality and bleeding risks in 1125 patients with COVID-19: relationship with anticoagulation status. Aging (Albany NY). 2021;13(7):9225–9242. doi:10.18632/aging.202769

20. Moores LK, Tritschler T, Brosnahan S, et al. Prevention, Diagnosis, and Treatment of VTE in Patients With Coronavirus Disease 2019: CHEST Guideline and Expert Panel Report. Chest. 2020;158(3):1143–1163. doi:10.1016/j.chest.2020.05.559

21. COVID-19 rapid guideline: reducing the risk of venous thromboembolism in over 16s with COVID-19. London: : National Institute for Health and Care Excellence (UK) 2020. http://www.ncbi.nlm.nih.gov/books/NBK566720/ (accessed 20 Apr 2021)

22. Antithrombotic Therapy to Ameliorate Complications of COVID-19 (ATTACC) - Full Text View - ClinicalTrials.gov. https://clinicaltrials.gov/ct2/show/NCT04372589 (accessed 9 Jan2021).

23. Tang N, Bai H, Chen X, Gong J, Li D, Sun Z. Anticoagulant treatment is associated with decreased mortality in severe coronavirus disease 2019 patients with coagulopathy. J Thromb Haemost. 2020;18(5):1094–1099. doi:10.1111/jth.14817

24. Paranjpe I, Fuster V, Lala A, et al. Association of Treatment Dose Anticoagulation With In-Hospital Survival Among Hospitalized Patients With COVID-19. J Am Coll Cardiol. 2020;76(1):122–124. doi:10.1016/j.jacc.2020.05.001

25. Protective Effect of Aspirin on COVID-19 Patients - Full Text View - ClinicalTrials.gov. https://clinicaltrials.gov/ct2/show/NCT04365309 (accessed 9 Jan2021).

26. Pavoni V, Gianesello L, Pazzi M, Stera C, Meconi T, Frigieri FC. Venous thromboembolism and bleeding in critically ill COVID-19 patients treated with higher than standard low molecular weight heparin doses and aspirin: A call to action. Thromb Res. 2020;196:313–317. doi:10.1016/j.thromres.2020.09.013

27. Cuker A, Tseng EK, Nieuwlaat R, et al. American Society of Hematology 2021 guidelines on the use of anticoagulation for thromboprophylaxis in patients with COVID-19. Blood Adv. 2021;5(3):872–888. doi:10.1182/bloodadvances.2020003763

28. Siegal DM, Barnes GD, Langlois NJ, et al. A toolkit for the collection of thrombosis-related data elements in COVID-19 clinical studies. Blood Adv. 2020;4(24):6259–6273. doi:10.1182/bloodadvances.2020003269

29. Schulman S, Angerås U, Bergqvist D, et al. Definition of major bleeding in clinical investigations of antihemostatic medicinal products in surgical patients. J Thromb Haemost. 2010;8(1):202–204. doi:10.1111/j.1538-7836.2009.03678.x

30. Kaatz S, Ahmad D, Spyropoulos AC, Schulman S; Subcommittee on Control of Anticoagulation. Definition of clinically relevant non-major bleeding in studies of anticoagulants in atrial fibrillation and venous thromboembolic disease in non-surgical patients: communication from the SSC of the ISTH. J Thromb Haemost. 2015;13(11):2119–2126. doi:10.1111/jth.13140

31. COVID-19 Treatment Guidelines Panel. Coronavirus Disease 2019 (COVID-19) Treatment Guidelines. National Institutes of Health. Available at https://www.covid19treatmentguidelines.nih.gov/. accessed [6/14/2021

32. Roomi SS, Saddique M, Ullah W, et al. Anticoagulation in COVID-19: a single-center retrospective study. J Community Hosp Intern Med Perspect. 2021;11(1):17-22. Published 2021 Jan 26. doi:10.1080/20009666.2020.1835297

33. Therapeutic Anticoagulation in Non-Critically Ill Patients with Covid-19 The ATTACC, ACTIV-4a, and REMAP-CAP Investigators, Patrick R. Lawler, Ewan C. Goligher, Jeffrey S. Berger, Matthew D. Neal, Bryan J. McVerry, Jose C. Nicolau, Michelle N. Gong, Marc Carrier, Robert S. Rosenson, Harmony R. Reynolds, Alexis F. Turgeon, Jorge Escobedo, David T. Huang, Charlotte Ann Bradbury, Brett L. Houston, Lucy Z. Kornblith, Anand Kumar, Susan R. Kahn, Mary Cushman, Zoe McQuilten, Arthur S. Slutsky, Keri S. Kim, Anthony C. Gordon, Bridget-Anne Kirwan, Maria M. Brooks, Alisa M. Higgins, Roger J. Lewis, Elizabeth Lorenzi, Scott M. Berry, Lindsay R. Berry, Derek C. Angus, Colin J. McArthur, Steven A. Webb, Michael E. Farkouh, Judith S. Hochman, Ryan Zarychanski medRxiv 2021.05.13.21256846; doi: https://doi.org/10.1101/2021.05.13.21256846

34. Khider L, Gendron N, Goudot G, et al. Curative anticoagulation prevents endothelial lesion in COVID-19 patients. J Thromb Haemost. 2020;18(9):2391–2399. doi:10.1111/jth.14968

35. Chocron R, Galand V, Cellier J, et al. Anticoagulation Before Hospitalization Is a Potential Protective Factor for COVID-19: Insight From a French Multicenter Cohort Study. J Am Heart Assoc. 2021;10(8):e018624. doi:10.1161/JAHA.120.018624

36. RECOVERY Collaborative Group, Horby P, Lim WS, et al. Dexamethasone in Hospitalized Patients with Covid-19. N Engl J Med. 2021;384(8):693–704. doi:10.1056/NEJMoa2021436

37. Bansal V, Mahapure KS, Bhurwal A, et al. Mortality Benefit of Remdesivir in COVID-19: A Systematic Review and Meta-Analysis. Front Med (Lausanne). 2021;7:606429. Published 2021 Jan 27. doi:10.3389/fmed.2020.606429

38. RECOVERY Collaborative Group. Tocilizumab in patients admitted to hospital with COVID-19 (RECOVERY): a randomised, controlled, open-label, platform trial. Lancet. 2021;397(10285):1637–1645. doi:10.1016/S0140-6736(21)00676-0

39. Laguna-Goya R, Utrero-Rico A, Talayero P, et al. IL-6-based mortality risk model for hospitalized patients with COVID-19. J Allergy Clin Immunol. 2020;146(4):799-807.e9. doi:10.1016/j.jaci.2020.07.009

40. Panka BA, de Grooth HJ, Spoelstra-de Man AM, Looney MR, Tuinman PR. Prevention or Treatment of Ards With Aspirin: A Review of Preclinical Models and Meta-Analysis of Clinical Studies. Shock. 2017;47(1):13–21. doi:10.1097/SHK.0000000000000745

41. Chow JH, Khanna AK, Kethireddy S, et al. Aspirin Use Is Associated With Decreased Mechanical Ventilation, Intensive Care Unit Admission, and In-Hospital Mortality in Hospitalized Patients With Coronavirus Disease 2019. Anesth Analg. 2021;132(4):930–941. doi:10.1213/ANE.0000000000005292

42. Viecca M, Radovanovic D, Forleo GB, Santus P. Enhanced platelet inhibition treatment improves hypoxemia in patients with severe Covid-19 and hypercoagulability. A case control, proof of concept study. Pharmacol Res. 2020;158:104950. doi:10.1016/j.phrs.2020.104950

43. Paranjpe I, Fuster V, Lala A, et al. Association of Treatment Dose Anticoagulation With In-Hospital Survival Among Hospitalized Patients With COVID-19. J Am Coll Cardiol. 2020;76(1):122–124. doi:10.1016/j.jacc.2020.05.001

