## Supplementary material for "Combined Anticoagulant and antiplatelet therapy is associated with an improved outcome in hospitalized COVID-19 patients: a propensity matched cohort study": supplementary material july 14.docx

**Table 3:** **Mortality model: 2 by 2**

|  | ProphAC | TherAC | ProphAC+AP | TherAC+AP |
| --- | --- | --- | --- | --- |
| ProphAC |  | 0.075 | 0.770 | 0.003 |
| TherAC | 0.075 |  | 0.429 | 0.809 |
| ProphAC+AP | 0.770 | 0.429 |  | 0.039 |
| TherAC+AP | 0.003 | 0.809 | 0.039 |  |

**Table 4: Propensity Score for anticoagulation(a) and antiplatelet prescriptioin(b)**

(a) **Propensity score for anticoagulation prescription**

| **Variables in the Equation** | | | | | | | | | |
| --- | --- | --- | --- | --- | --- | --- | --- | --- | --- |
|  | | B | S.E. | Wald | df | Sig. | Exp(B) | 95% C.I.for EXP(B) | |
|  |  |  |  |  |  |  |  | Lower | Upper |
|  | Admission HR | -.028 | .010 | 7.658 | 1 | .006 | .973 | .954 | .992 |
|  | CHF | 1.534 | .712 | 4.638 | 1 | .031 | 4.637 | 1.148 | 18.731 |
| a. Variable(s) entered on step 1: Age, Gender, Smoker, Weight, AdmissionHR, AdmissionTemperature, AdmissionOxygenSaturation, HTN, Dyslipedemia, CHF, Cancer, DM, HistoryBleeding, Liverdisease, KidneyDisease, COPD, CAD, PreviousVTE, PlateletCount, SMEAN(ddimers), SMEAN(CRP), SMEAN(Interleukin6), AnticoagulationAdmission, SMEAN(Fibrinogen), SMEAN(Troponin). | | | | | | | | | |

(b) **Propensity scores for antiplatelet prescription**

| **Variables in the Equation** | | | | | | | | | |
| --- | --- | --- | --- | --- | --- | --- | --- | --- | --- |
|  | | B | S.E. | Wald | df | Sig. | Exp(B) | 95% C.I.for EXP(B) | |
|  |  |  |  |  |  |  |  | Lower | Upper |
|  | Gender | -.910 | .348 | 6.820 | 1 | .009 | .403 | .203 | .797 |
|  | Smoker | -.667 | .370 | 3.257 | 1 | .071 | .513 | .249 | 1.059 |
|  | AdmissionTemperature | -.382 | .160 | 5.680 | 1 | .017 | .682 | .498 | .934 |
|  | COPD | 5.439 | 3.007 | 3.271 | 1 | .070 | 230.290 | .635 | 83580.632 |
|  | SMEAN(ddimers) | -.530 | .148 | 12.782 | 1 | .000 | .589 | .440 | .787 |
|  | AntiPAdmission | 2.772 | .720 | 14.838 | 1 | .000 | 15.985 | 3.902 | 65.495 |
|  | Constant | 9.237 | 6.494 | 2.023 | 1 | .155 | 10271.108 |  |  |
| a. Variable(s) entered on step 1: Age, Gender, Smoker, Weight, AdmissionHR, AdmissionTemperature, AdmissionOxygenSaturation, HTN, Dyslipedemia, CHF, Cancer, DM, HistoryBleeding, Liverdisease, KidneyDisease, COPD, CAD, PreviousVTE, PlateletCount, SMEAN(ddimers), SMEAN(CRP), SMEAN(Interleukin6), AntiPAdmission, SMEAN(Fibrinogen), SMEAN(Troponin). | | | | | | | | | |
